# Optimal Size of COVID-19 Testing Pools

**DOI:** 10.1101/2020.07.20.20158436

**Authors:** Jon Cohen

## Abstract

This research note investigates the optimal size of pools for pooled, COVID-19 testing when positive pools will be followed up by individual tests of pool members. Formulae for the optimum are derived and provided. The analysis indicates that

- optimal pool sizes are unlikely to exceed about 20 individuals in realistic situations,
- optimal pool size is influenced by prevalence in the population and the extent to which infection is clustered within the population, and,
- pools are most efficiently comprised of people with homogeneous risk, with heterogeneity across pools.

## Optimal size and composition of COVID-19 testing pools

As the country returns to work from closures associated with Covid-19, employers are seeking mechanisms to help keep their workers and communities safe and healthy. While it is widely understood that testing is necessary, the costs can prove prohibitive when applied to large number of individuals.

Approaches have been proposed to pool samples taken from multiple individuals and test these pooled samples with a single test. One of the most expensive parts of the test is the processing, which uses specialized reagents to detect the virus within the sample. The tests used for COVID-19 are very sensitive and can detect a very diluted concentration of virus. Pooling involves taking samples from multiple people and processing them together using a single batch of reagents. This drastically reduces cost, but provides more limited information. Results provide information about whether any of the people included in the pool is COVID-19 positive.

Pooled testing, the process of aggregating individual samples for processing with a single batch of reagents, can provide the basis for cost-effective COVID-19 screening for entire workforces. Testing would occur in three steps:

- Divide the site population (e.g., workforce, students) into pools, probably based on the proximity of their work-spaces;
- Periodically test the pools to identify any pools that include at least one infected individual; and
- Individually test every member of any pool that yields a positive result.

This note evaluates the optimal size of the testing pools, and identifies factors that can minimize the total number of tests required. Before getting into the technical details, I offer a simple example. Consider the example of a workplace with 100 workers with an (as yet unknown) infection rate of one percent. At a price of $150 per test, testing every worker would cost $15,000 in total, and that cost would recur at regular intervals. Now, imagine a regime in which groups of 10 workers are tested with a single, pooled test. Only one or two pools are likely to include COVID-positive individuals, so the total number of tests would be between 10 (if no pools contain positive individuals) and 30 (if two of the pools contain positive individuals). The total cost ranges from $1,500 to an upper limit of $4,500, with an expectation of $3,000, or one fifth of the cost of testing everyone. Effectively, this approach protects a workforce of 100 people for an expected cost of about $30 per person for the full workforce, compared to $150 per person if individual tests are applied universally.

The testing plan proposed above covers the entire population. The goal, then, is to get the coverage at the lowest cost. We achieve that by minimizing the total number of tests. Let the total number of tests required (*T*) be

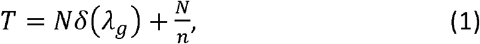

where *T* represents the total number of tests, *n* is the number of workers included in each pool of tests, and N is the total size of the workforce (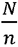 is the number of pools in the population, each requiring one test).

The function *δ* (*λ*_*g*_) represents the probability that a group includes at least one COVID-positive person. Modeling this with a mixture of Poisson distributions representing the testing pools, recognizing that each pool may have a different prevalence of COVID-19, but combined they have an expected value of *nr*.

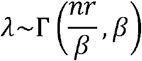

where *E* (*λ*) = *nr*, where *r* is the (initially unknown) rate of infection in the target population. *β* is a scale parameter and reflect heterogeneity in infection rates across groups. Of course, the infection rate for each group is unknown before testing, so we must integrate over the distribution of *λ* to get an expected value. The critical rate, *r*, is also unknown so we must begin with a reasonable estimate, and refine the value as data accumulate. The probability of a pool testing positive because one or more member of the pool is positive is derived by integrating over the *λ* distribution,

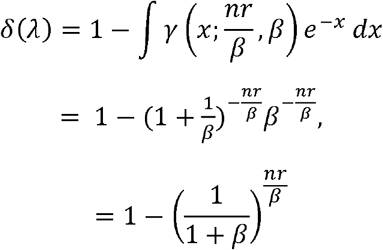

where *γ*(.) is the gamma probability density function and the second term is the Poisson probability mass function at zero (the probability that the group has no infected individuals).

The optimum group size minimizes the number of tests

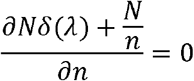

So

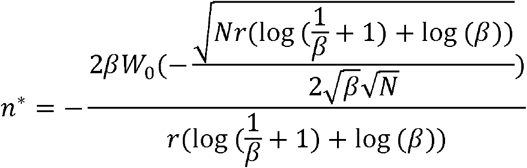

Where *w*_0_is the principal branch of the Lambert W function, for which convenient approximations exist (e.g., de Bruijn, 1981; Weisstein, 2020). Noting that N > 0, this simplifies further to

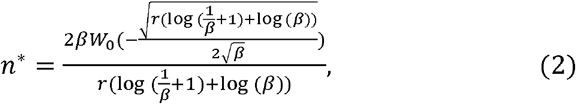

which is independent of *N*, the total number of employees. Tables 1–3 present the optimal testing pool sizes and expected number of tests required to cover a population of 1,000 workers. The size of the workforce is arbitrary in these examples, and the costs and associated calculations scale linearly with workforce size, as implied in Equation 2. As a basis for comparison, recognize that each round of testing would cost $150,000 to cover all employees. As Tables 1–3 show, our hybrid model of pooled testing and individual testing of those in COVID-19 positive pools can reduce the cost of testing the full workforce by an order of magnitude in some cases.

**Table 1:**
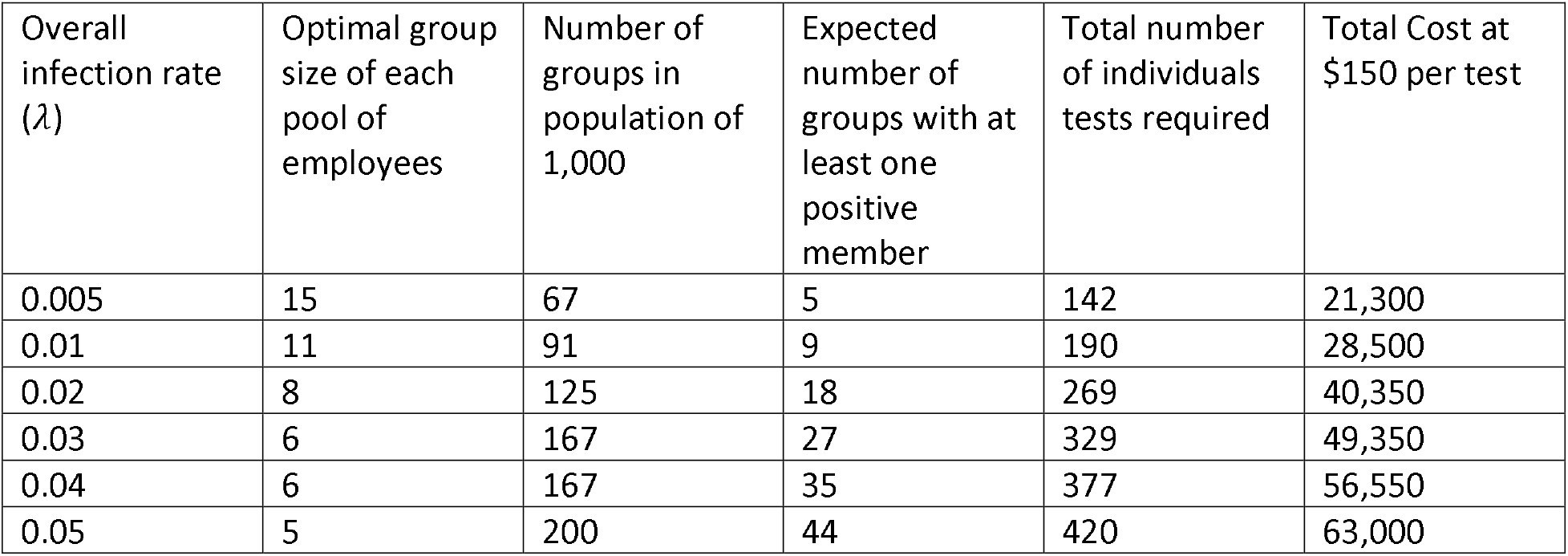
Optimal testing groups sizes, numbers of tests, and costs given the overall infection rate with *β* = .01 (virtually no variation across groups)

Each table presents sample values for underlying infection rates ranging from 0.5% to 5.0%. Table 1 presents a case with virtually homogenous infection rates across groups. Table 2 introduces moderate variation in the infection rates across groups, and Table 3 introduces substantial variation in the infection rates across groups.

**Table 2:** Optimal testing groups sizes, numbers of tests, and costs given the overall infection rate with *β* = .1 (moderate variation across groups)

| Overall infection rate ($\lambda$) | Optimal group size | Number of groups in population of 1,000 | Expected number of groups with at least one positive member | Total number of tests required | Total Cost at \$150 per test |
| --- | --- | --- | --- | --- | --- |
| 0.005 | 18 | 56 | 3 | 110 | 16,500 |
| 0.01 | 13 | 77 | 7 | 168 | 25,200 |
| 0.02 | 9 | 112 | 13 | 229 | 34,350 |
| 0.03 | 7 | 143 | 19 | 276 | 41,400 |
| 0.04 | 7 | 143 | 25 | 318 | 47,700 |
| 0.05 | 6 | 167 | 31 | 353 | 52,950 |

**Table 3:** Optimal testing groups sizes, numbers of tests, and costs given the overall infection rate with *β* = 2 (substantial variation across groups)

| Overall infection rate ($\lambda$) | Optimal group size | Number of groups in population of 1,000 | Expected number of groups with at least one positive member | Total number of tests required | Total Cost at \$150 per test |
| --- | --- | --- | --- | --- | --- |
| 0.005 | 20 | 50 | 3 | 110 | 16,500 |
| 0.01 | 14 | 72 | 5 | 142 | 21,300 |
| 0.02 | 10 | 100 | 10 | 200 | 30,000 |
| 0.03 | 8 | 125 | 16 | 253 | 37,950 |
| 0.04 | 7 | 143 | 20 | 283 | 42,450 |
| 0.05 | 7 | 143 | 25 | 318 | 47,700 |

Notice that as we increase the between-group variation in infection rates, the optimal group sizes increase and the total number of tests required decrease, along with their costs. This dynamic is illustrated below in Figure 1.

**Figure 1:**
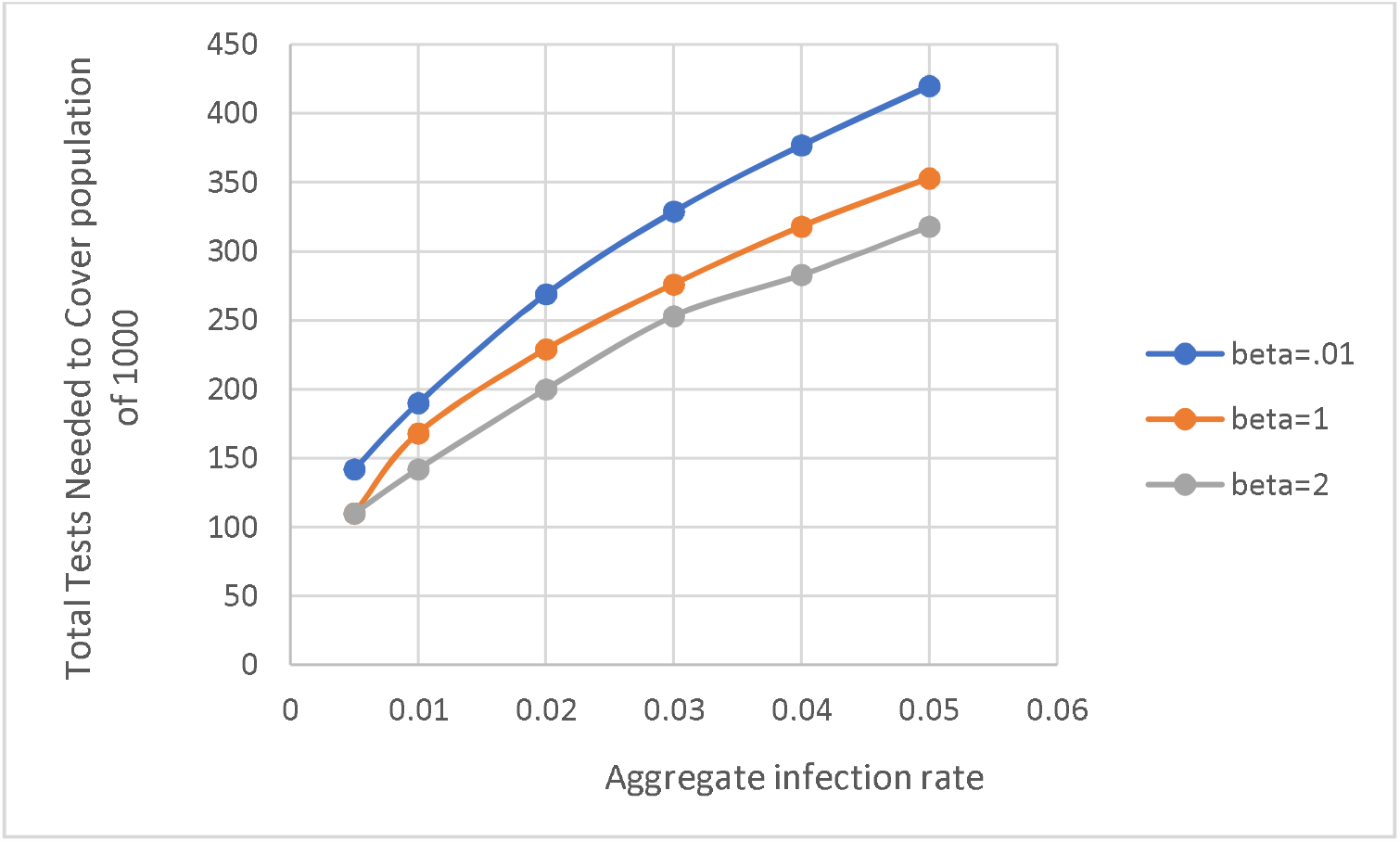
Total tests needed by aggregate infection rate and homogeneity of testing groups.

Figure 1 pulls together data from the tables to illustrate the impact of COVID-19 prevalence and within-group contagion on the total number of tests needed. The top (blue) line shows the number of tests needed if groups are formed at random. With random assigned to groups an infection rate of 3 percent would require nearly 350 tests. If the groups are formed so that the risk is more similar within group and more variable across groups (the bottom, grey line), about 250 tests can cover the population. At $150 per test, this represents a substantial savings.

Initial testing would have to rely on available best-guess estimates of infection rates and cross-group heterogeneity. As data is gathered these estimates can be updated to reflect workplace-specific factors. One thing made clear by Figure 1 is that heterogeneous risk across pools increases the efficiency of testing. Given how contagion works, forming groups according to the physical proximity of the individuals to each other should help increase this factor, which will increase the efficiency of the group testing.

As tests are conducted and data gathered, the data can be used to estimate the actual values of the various parameters. We can model the group-specific infection rates as a function of workplace-specific factors such as worksite, office layout and physical job requirements, and this information can be used to stratify the sample, optimizing group sizes to optimize testing resources. A hybrid testing model makes it possible for an organization to protect their community population and workforce and their balance sheets at the same time.

## Data Availability

This is an analytic piece, so it is not a data analysis

